# Cancer-Type Specific Prognostic Impact of Concurrent TP53 and KRAS Alterations: A Multi-Cohort Genomic Analysis

**DOI:** 10.64898/2026.03.29.26349383

**Authors:** Pan Guangchao

## Abstract

**Background:** The tumor suppressor gene *TP53* and the oncogene *KRAS* are among the most frequently altered core drivers in human malignancies. Although they cooperatively regulate critical biological processes, the prognostic impact of their co-alterations remains poorly defined and exhibits striking inconsistency across different cancer types.

**Methods:** We comprehensively analyzed genomic and clinical data from multi-cancer cohorts sourced from the cBioPortal database and The Cancer Genome Atlas (TCGA). Genetic alterations, including sequence variations and copy number alterations (CNAs), were classified for *TP53* and *KRAS*. Patients were stratified into four subgroups based on individual or combined alteration status. Survival analyses were performed using Kaplan-Meier methods. Integrated multi-omics analyses were conducted to assess the relationship between genetic alterations and mRNA/protein expression, and to characterize co-occurring genetic events and their prognostic implications.

**Results:** Patients harboring concurrent *TP53* and *KRAS* alterations exhibited significantly shorter overall survival in pancreatic cancer, colorectal cancer, and ampullary carcinoma, but surprisingly demonstrated the longest survival in gastric cancer. Distinct *KRAS* mutation subtype distributions were observed across cancer types: G12D/G12V predominated in pancreatic and colorectal cancers, G12C in non-small cell lung cancer, and G13D in gastric cancer, with copy number alterations representing a substantial proportion of *KRAS* alterations in gastric and lung cancers. Multi-omics analysis revealed a lack of concordance between genetic alterations and mRNA/protein expression, indicating that mutation status alone does not reliably reflect downstream molecular changes. Concurrent genetic events displayed striking cancer-type specificity: *CDKN2A* alterations frequently co-occurred with *TP53*/*KRAS* double alterations in pancreatic cancer and were associated with worse prognosis, whereas *APC* mutations co-occurred in colorectal cancer and correlated with improved survival. Integrated analysis further demonstrated that *KRAS*^altered^/*TP53*^altered^ patients were highly enriched in pancreatic, colorectal, and lung cancers, each exhibiting unique background genomic landscapes.

**Conclusions:** The prognostic significance of *TP53* and *KRAS* alterations is profoundly cancer-type specific, driven by differences in mutation subtype distribution, copy number alteration patterns, co-occurring genetic events, and the discordance between genotype and functional expression. These findings challenge the simplistic view of dual-gene alterations as universal markers of poor prognosis and underscore the necessity of incorporating cancer-specific molecular contexts into prognostic models and precision oncology strategies.

## Introduction

TP53 is one of the most frequently mutated genes which alters in approximately 50% human cancers^1,2^. These mutations confer a broad range of functional consequences, including the loss of tumor suppressor activity, dominant-negative effects, and gain-of-oncogenic functions, which collectively drive tumor initiation, progression, and therapeutic resistance^3,4^. The copy number variations (CNVs) such as monoallelic or biallelic deletions of the p53 locus are also recurrent events in diverse malignancies, often cooperating with mutations^5,6^.. Moreover, the temporal order of p53 alterations relative to other driver mutations—and their interplay with concurrent genomic events—profoundly shapes tumor evolution and patient outcomes^3,7,8^. Despite considerable progress, a comprehensive understanding of the multifaceted roles of p53 mutations and copy number variations in cancer biology remains elusive.

KRAS amplifications and mutations exert their oncogenic effects by increasing the levels of active KRAS-GTP, leading to constitutive activation of downstream signaling pathways^9^. The prevalence of KRAS mutations is notably high in certain types of cancer. However, the distribution of specific KRAS mutation subtypes varies across cancer types: G12D predominates in PDAC^10,11^, whereas G12C is more common in lung adenocarcinoma^12^.

Distinct KRAS mutation subtypes are associated with varying patient prognoses^13^. Furthermore, the therapeutic outcomes and prognosis of KRAS-driven tumors are influenced by co-occurring genomic alterations. For instance, in KRAS-mutant lung adenocarcinoma, concurrent STK11/LKB1 or KEAP1 mutations confer primary resistance to PD-1 inhibitors and are associated with poorer outcomes^14,15^.

Notably, a solitary KRAS G12D mutation is insufficient to reliably induce pancreatic ductal adenocarcinoma (PDAC)^16,17^. Studies in mouse models have demonstrated that KRAS G12D must be combined with inactivation of tumor suppressor genes such as TP53 or CDKN2A to efficiently drive pancreatic cancer progression^16,18–20^. Indeed, approximately 50-75% of PDAC patients harbor concurrent TP53 mutations, and this cooperative effect significantly accelerates tumor initiation and aggressiveness^21,22^.

Mutations in KRAS and TP53 are two pivotal genetic alterations driving cancer initiation and progression. When both occur, their synergistic effect not only exacerbates the malignant phenotype but also profoundly impacts clinical prognosis and therapeutic response. For instance, in patients with colorectal liver metastases, RAS/TP53 co-mutation has been identified as an independent predictor of shorter overall survival^23^. In non-small cell lung cancer (NSCLC), this co-mutation pattern has emerged as a potential predictive biomarker for benefit from immunotherapy^24,25^. Furthermore, in PDAC, the presence of both mutations is associated with poor differentiation, increased lymph node involvement, and reduced survival following chemotherapy^26,27^.

These findings reveal that the co-occurrence of KRAS and TP53 mutations is not merely additive but involves complex molecular crosstalk that shapes a distinct and aggressive tumor biology. Therefore, there is an urgent need to re-evaluate such combined mutations and to explore potential pathological mechanisms.

## Results

### Prognosis differs in combination of *TP53* and *KRAS* genetic alterations across different cancer types

The tumor suppressor gene *TP53* and the oncogene *KRAS* are among the most frequently altered core drivers in human malignancies. They cooperatively regulate critical biological processes. However, the prognostic impact of their co-alterations remains poorly defined and inconsistent across cancer entities.

Based on comprehensive multi-cancer datasets from the cBioPortal database, we classified both copy number alterations (CNA) and sequence variations including single nucleotide variants (SNVs), insertions and deletions as genetic events. We stratified patients into four subgroups based on individual or combined alterations of *TP53* and *KRAS* and performed survival analyses.

Patients harboring concurrent *TP53* and *KRAS* (*KRAS*^*altered*^/*TP53*^*altered*^) alterations exhibited significantly shorter overall survival (OS) in pancreatic cancer (PC), colorectal cancer (CRC), and ampullary carcinoma relative to the remaining subgroups (Figure 1A–C). Meanwhile, the *KRAS*-altered/*TP53*-wild-type (*KRAS*^*altered*^/*TP53*^*wt*^) subgroup predicted inferior OS in non-small cell lung cancer (NSCLC) and gastric cancer (GC), whereas it was associated with favorable survival outcomes in appendiceal cancer and endometrial cancer (EMC) (Figure 1D–G).

**Figure 1.**
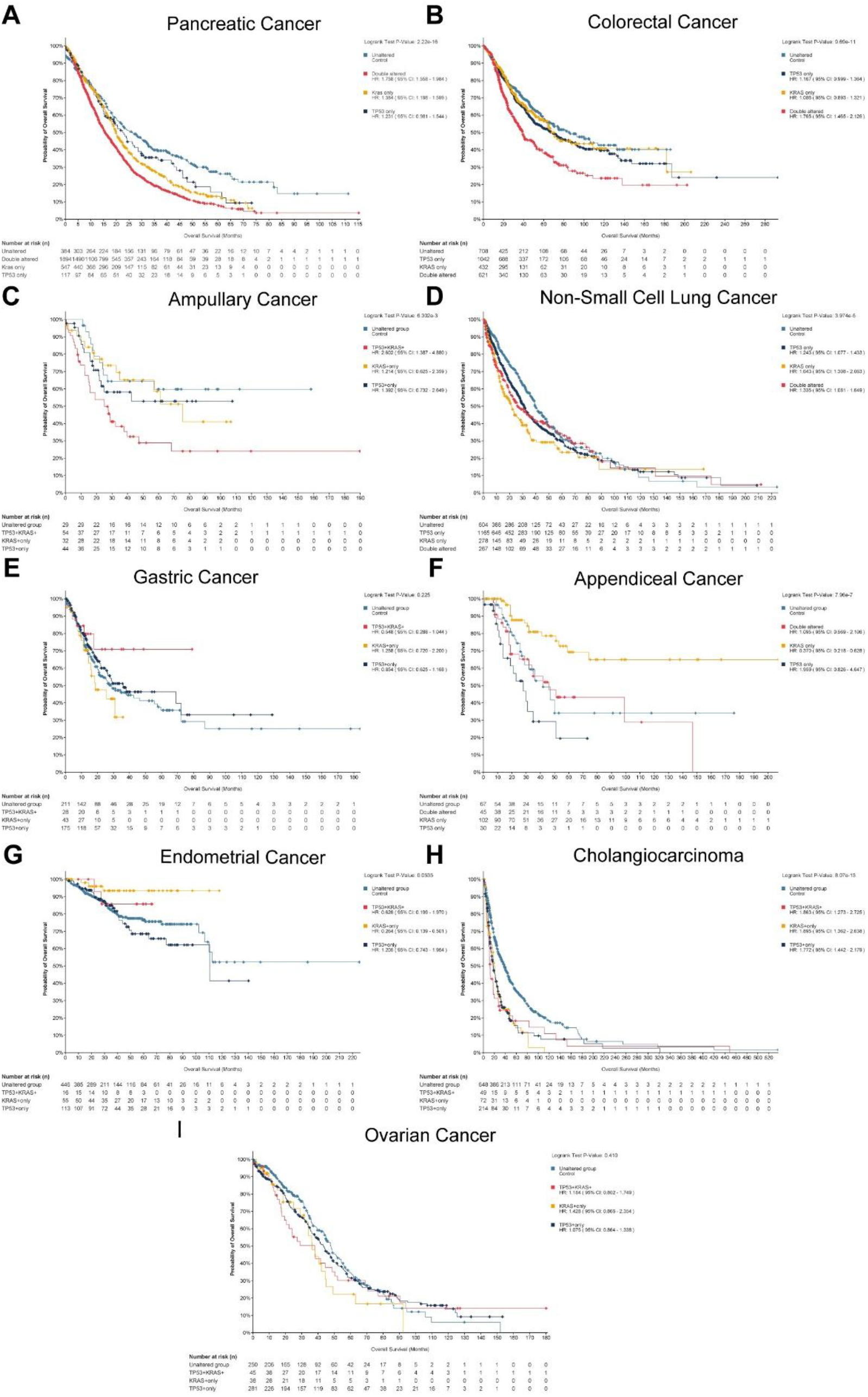
Overall survival analysis stratified by *TP53* and *KRAS* alteration status across different cancer types. Non-redundant datasets from the cBioPortal database were analyzed. Kaplan–Meier survival curves comparing the four subgroups (*KRAS*^altered^/*TP53*^altered^, *KRAS*^altered^/*TP53*^wt^, *KRAS*^wt^/*TP53*^altered^, and double wild-type) are shown for **(A)** PC, **(B)** CRC, **(C)** ampullary carcinoma, **(D)** NSCLC, **(E)** GC, **(F)** appendiceal cancer, **(G)** EMC, **(H)** cholangiocarcinoma, and **(I)** ovarian cancer.

Notably, *KRAS*^*altered*^/*TP53*^*altered*^ cases displayed prolonged OS in GC and the longest central nervous system progression-free survival (CNS PFS) in NSCLC (Figure 1E, Supplementary Figure 1A–B). Collectively, the prognostic significance of single and combined *TP53*/*KRAS* alterations lacks universal applicability and exhibits striking tumor-type specificity, suggesting that gene stratification based on these two genes confers important clinical prediction across different cancer types.

**Supplementary Figure 1.**
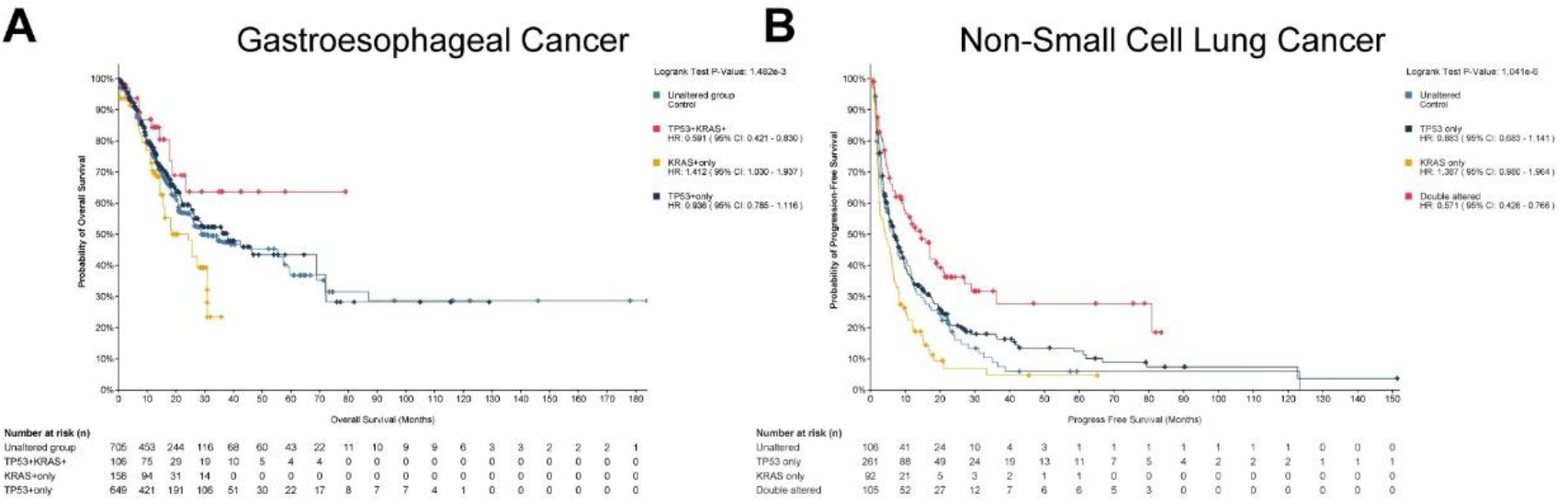
**(A)** Overall survival analysis in GC including all samples (allowing duplicate samples) from the cBioPortal database. **(B)** Central nervous system progression-free survival (CNS PFS) analysis in NSCLC.

### Genetic characteristics of *KRAS*^*altered*^/*TP53*^*altered*^ cases across distinct cancer types

To elucidate underlying molecular mechanisms for the aforementioned prognostic heterogeneity, we selected core cancer types with significantly divergent prognosis in the dual-gene alteration group as the key research subjects, including PC, CRC, ampullary carcinoma, GC and NSCLC (Figure 1). Meanwhile, mutations with a frequency below 5% were defined as low-frequency mutations to stratify the mutation distribution spectrum. *TP53* alterations were dominated by low-frequency point mutations, and only the R175H variant represented the core high-frequency mutation site. In contrast, *KRAS* alterations were mainly driven by gene mutations, with a negligible proportion of gene amplification events; notably, high-frequency *KRAS* amplification was only detected in GC and NSCLC (Figure 2A). Stratified analysis of KRAS mutation subtypes revealed that *KRAS* G12D and G12V mutations were predominantly enriched in pancreatic adenocarcinoma (PC), colorectal carcinoma (CRC), and ampullary carcinoma. In contrast, gene amplification emerged as the primary driver rather than point mutations in GC. Notably, the coexistence of *KRAS* G12C mutation and gene amplification represented the dominant molecular signature in NSCLC (Figure 2A).

**Figure 2.**
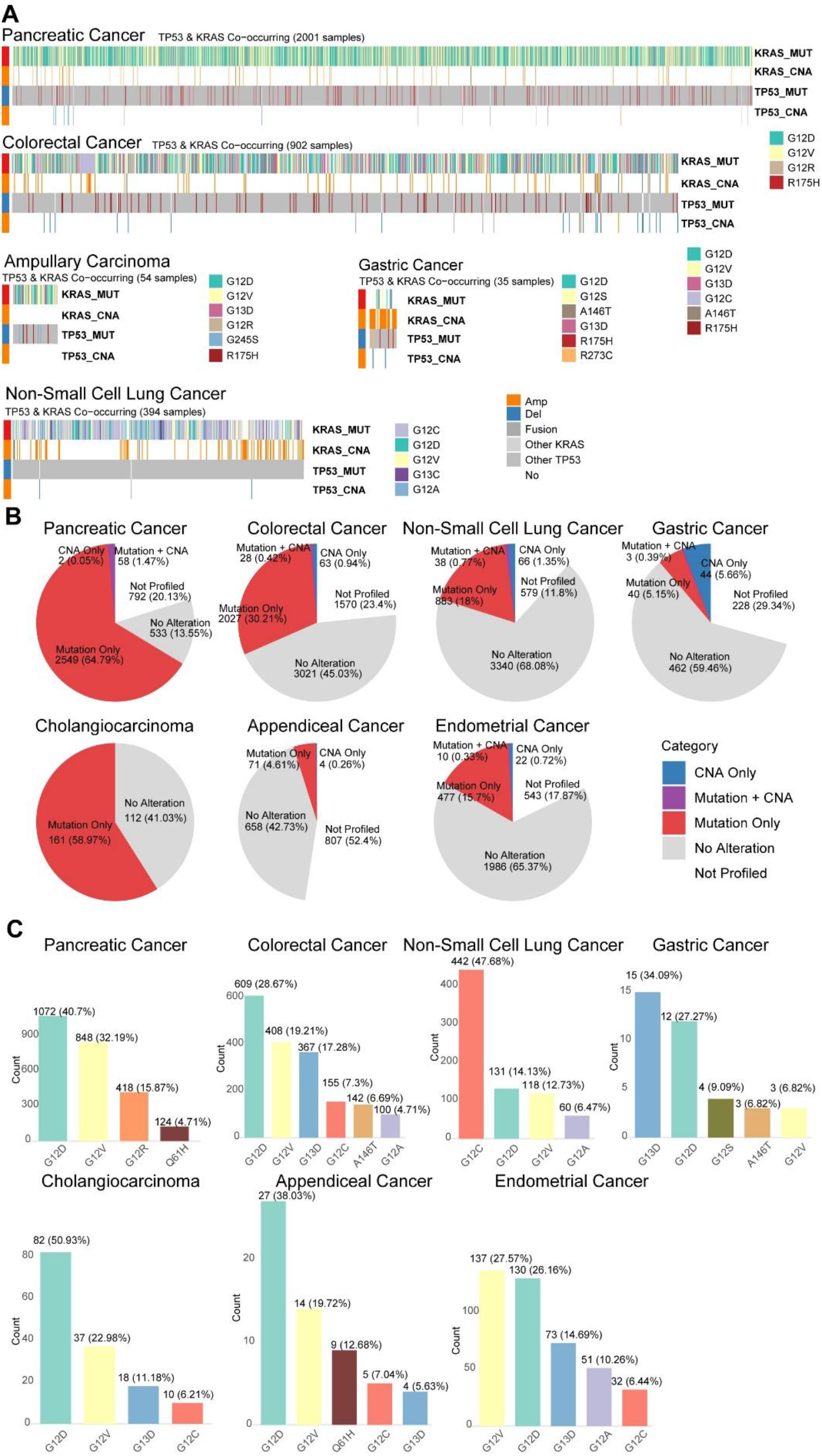
Landscape of *KRAS* alterations across cancer types. **(A)** Heatmap depicting the distribution of *KRAS* and *TP53* alteration types in double-alteration samples across five selected cancer types. **(B)** Proportions of different *KRAS* alteration types in seven cancer types. **(C)** Frequencies of high-frequency *KRAS* mutation subtypes across the same seven cancer types.

Furthermore, the *KRAS*^*altered*^/*TP53*^*wt*^ subgroup also exhibited distinct prognostic heterogeneity across cancer types (Figure 1). We further examined the landscape of KRAS alterations across these cancer types. The *KRAS*-altered/*TP53*-wt subgroup was associated with poorer clinical outcomes in PC, NSCLC, GC and cholangiocarcinoma, whereas it predicted favorable prognosis in appendiceal adenocarcinoma and EMC (Figure 1). However, the frequency of *KRAS* alterations differed markedly across cancer types: *KRAS* mutations accounted for 64% in PC and 58% in cholangiocarcinoma, representing the major driver events in these two cancer types. The proportion of *KRAS* alterations was lowest in appendiceal adenocarcinoma. In GC, despite the low alteration frequency, the proportion of CNAs was the highest, comprising more than half of all *KRAS* alterations (Figure 2B). Regarding the distribution of precise mutation subtypes, *KRAS* G12D and G12V variants predominated in all enrolled cancer types except NSCLC and GC; in contrast, the core mutation subtype was G12C in NSCLC and G13D in GC (Figure 2C).

Collectively, *KRAS* mutations represent core driver events in PC, cholangiocarcinoma and CRC, with G12D and G12V as the predominant mutational subtypes. This subgroup was associated with poor prognosis even in the presence of *TP53* alterations. In the other enrolled cancer types, by contrast, the proportion of *KRAS* alterations was markedly reduced (below 30%), and the predominant patterns of *KRAS* alterations in NSCLC and GC differed substantially from those in gastrointestinal tumors. While these patterns are not significantly associated with prognostic heterogeneity, they imply that cancer type specific genetic backgrounds may modulate the oncogenic activity of oncogenes.

### Prognostic Validation and Multi-Omics Analysis Using the TCGA Dataset

Although the integrated multi-cohort dataset included sufficient sample size, it lacked mRNA transcriptomic profiles, limiting investigations into gene-level associations with transcriptional and translational regulation. We therefore further incorporated the standard TCGA dataset, which harbors matched mRNA and protein expression data, for supplementary prognostic validation and integrated multi-omics analysis. Following integration of mRNA expression and protein quantification data from the TCGA cohort, *TP53* and *KRAS* alterations exhibited novel multi-omics associations: in samples with gene amplification, more than half displayed significantly upregulated mRNA expression; most samples with homozygous deletion showed markedly reduced mRNA levels. In contrast, mRNA expression abnormalities were rare in samples with gene mutations, indicating that mutational events exert weak effects at the transcriptional level (Figure 3A). These findings suggest that genetic alterations cannot be directly equated with expression changes in prognostic analysis. Notably, in CRC and endometrial carcinoma, a considerable proportion of samples showed isolated mRNA and protein expression abnormalities without detectable gene mutations or copy number variations. By contrast, such independent transcriptional and proteomic aberrations were much less in PC (Figure 3A).

**Figure 3.**
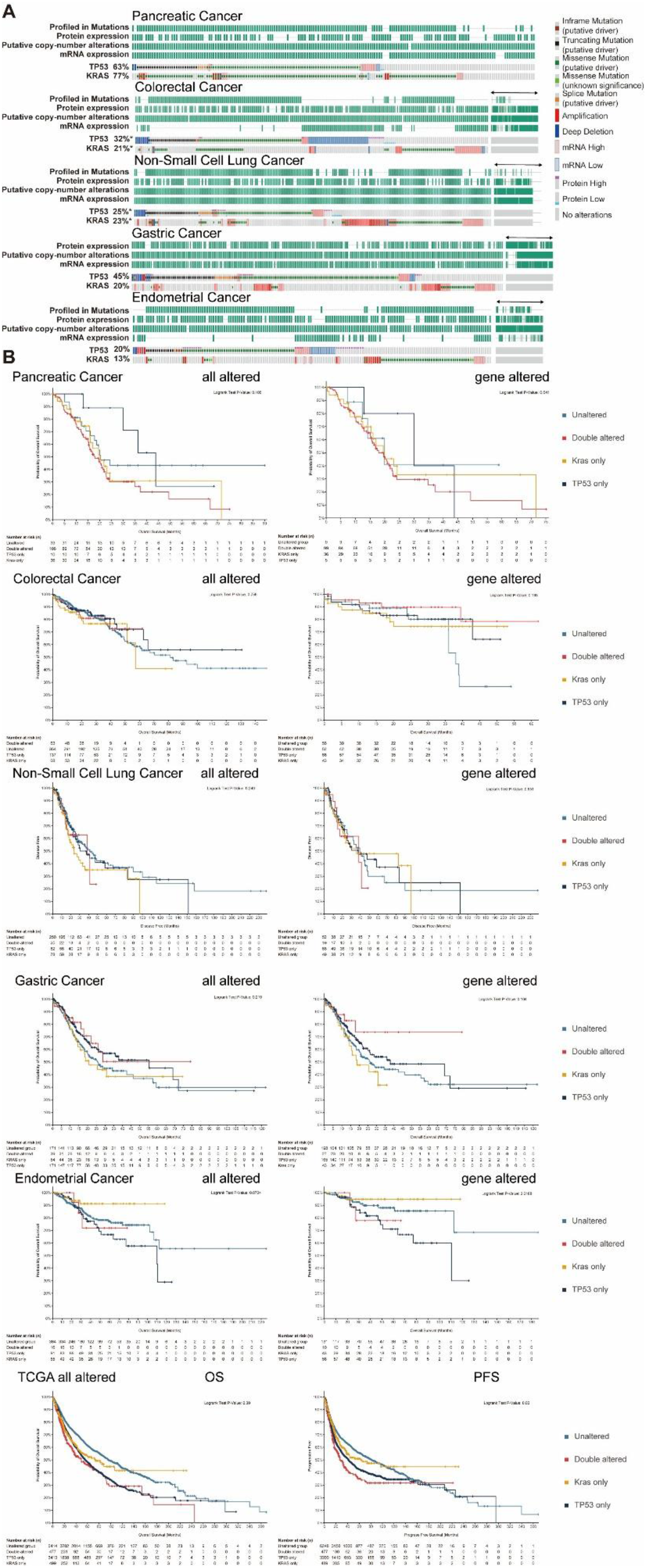
Multi-omics analysis and prognostic stratification in the TCGA cohort. **(A)** Landscape of *TP53* and *KRAS* alterations in five cancer types based on TCGA data, including mutation, copy number alteration, mRNA expression, and protein expression. **(B)** Prognostic analysis stratified by *TP53* and *KRAS* alteration status in individual TCGA cancer cohorts. **(C)** Overall survival analysis in the pan-cancer TCGA cohort stratified by the four *TP53*/*KRAS* subgroups.

**Supplementary Figure 2.**
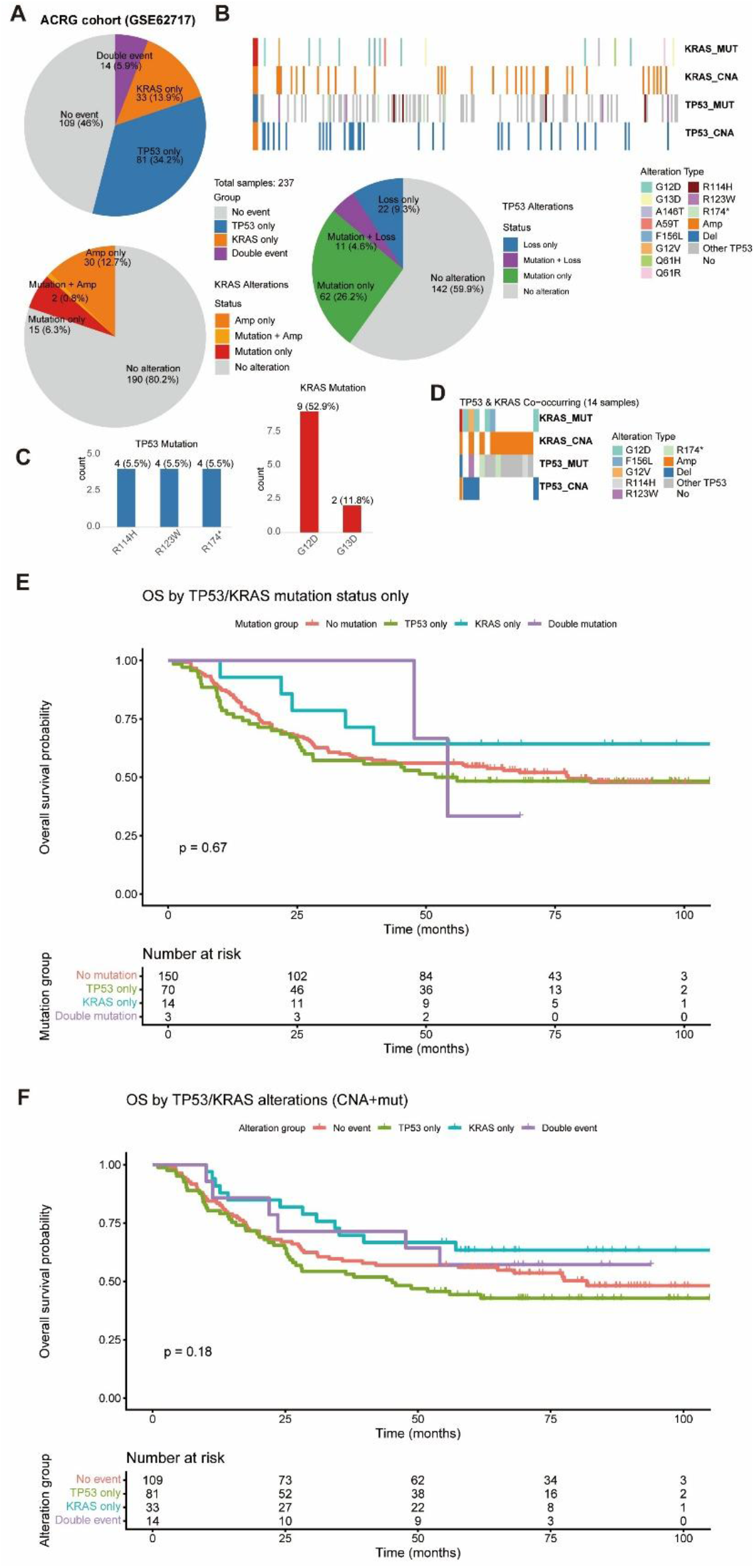
Comprehensive analysis of *TP53* and *KRAS* alterations in the ARCG dataset. **(A)** Pie chart showing the distribution of *TP53* and *KRAS* alteration events. **(B)** Heatmap of alteration profiles for both genes across all samples. **(C)** Bar plot of high-frequency mutations occurring in more than one sample. **(D)** Heatmap of co-alteration patterns in double-positive samples. **(E)** Prognostic analysis based on mutation status alone. **(F)** Prognostic analysis integrating both mutation and CNA status.

Based on the multi-omics characteristics, this study performed prognostic analyses on the single cancer-type specific datasets and the pan-cancer integrated cohort of TCGA. Two strategies were adopted for the analyses: grouping based solely on genetic alterations, and combined both mRNA and protein expression profiles. Notably, the sample size of each single cancer-type dataset was markedly smaller than the previous cohort, resulting in compromised statistical power. Only the prognostic differences based on genetic alteration grouping in EMC reached statistical significance (Figure 3B), confirming that *KRAS* alterations serve as a stable prognostic factor in this cancer type. In GC, the prognostic trend observed in the genetic alteration-only grouping remained consistent with that of the preliminary integrated cohort, whereas the combined grouping incorporating mRNA and protein expression profiles failed to replicate this trend (Figure 3B). This finding suggests that the prognostic model is primarily driven by genetic alterations rather than integrated multi-omics signatures.

Comprehensive analysis of the TCGA pan-cancer integrated cohort revealed that patients stratified into four distinct subgroups based on combined *KRAS* and *TP53* gene alteration status exhibited divergent prognostic profiles (Figure 3B). This observation suggests that concurrent gene alterations of *TP53* and *KRAS* do not exert a simple additive oncogenic effect, but rather confer complex and heterogeneous prognostic implications across cancer types.

Collectively, there was no significant consistent correlation between *TP53* and *KRAS* genetic alterations and their corresponding mRNA and protein expression levels. Meanwhile, prognostic stratification solely based on genetic alteration status is not applicable when incorporating mRNA and protein expression variations into the grouping model.

To further supplement the data, we incorporated the ACRG dataset. Analysis revealed that, compared with GC samples from the cBioPortal database, the ACRG cohort exhibited a higher frequency of *KRAS* amplification, with the G12D mutation being the most common and G13D the second most frequent *KRAS* mutation (Supplemental Figure 2A–C). Additionally, *TP53* mutations were no longer dominated by the R175H hotspot (Supplemental Figure 2D). In prognostic analyses based on mutation and copy number alteration stratification, the *KRAS*^*altered*^/*TP53*^*wt*^ subgroup showed favorable outcomes (Supplemental Figure 2E-F). These differences may suggest heterogeneity between Eastern and Western gastric cancer patient populations.

### Integrated Analysis of Concurrent Genetic Features and Stratified Prognosis in *KRAS*^*altered*^/*TP53*^*altered*^ Patients Across the Entire Cohort

To further explore the distinct genetic characteristics of *KRAS*^*altered*^/*TP53*^*altered*^ patients, this study performed a comprehensive pooled analysis of those samples within the entire study cohort. *KRAS*^*altered*^/*TP53*^*altered*^ samples were highly enriched in PC, CRC and NSCLC, with these three cancer types accounting for more than 80% population (Figure 4A). Subsequently, tumor mutational burden (TMB) analysis was performed on cancer types with more than 100 double-positive samples. A small subset of cases with high TMB was identified in CRC and NSCLC, whereas almost none were observed in breast cancer (Figure 4B).

**Figure 4.**
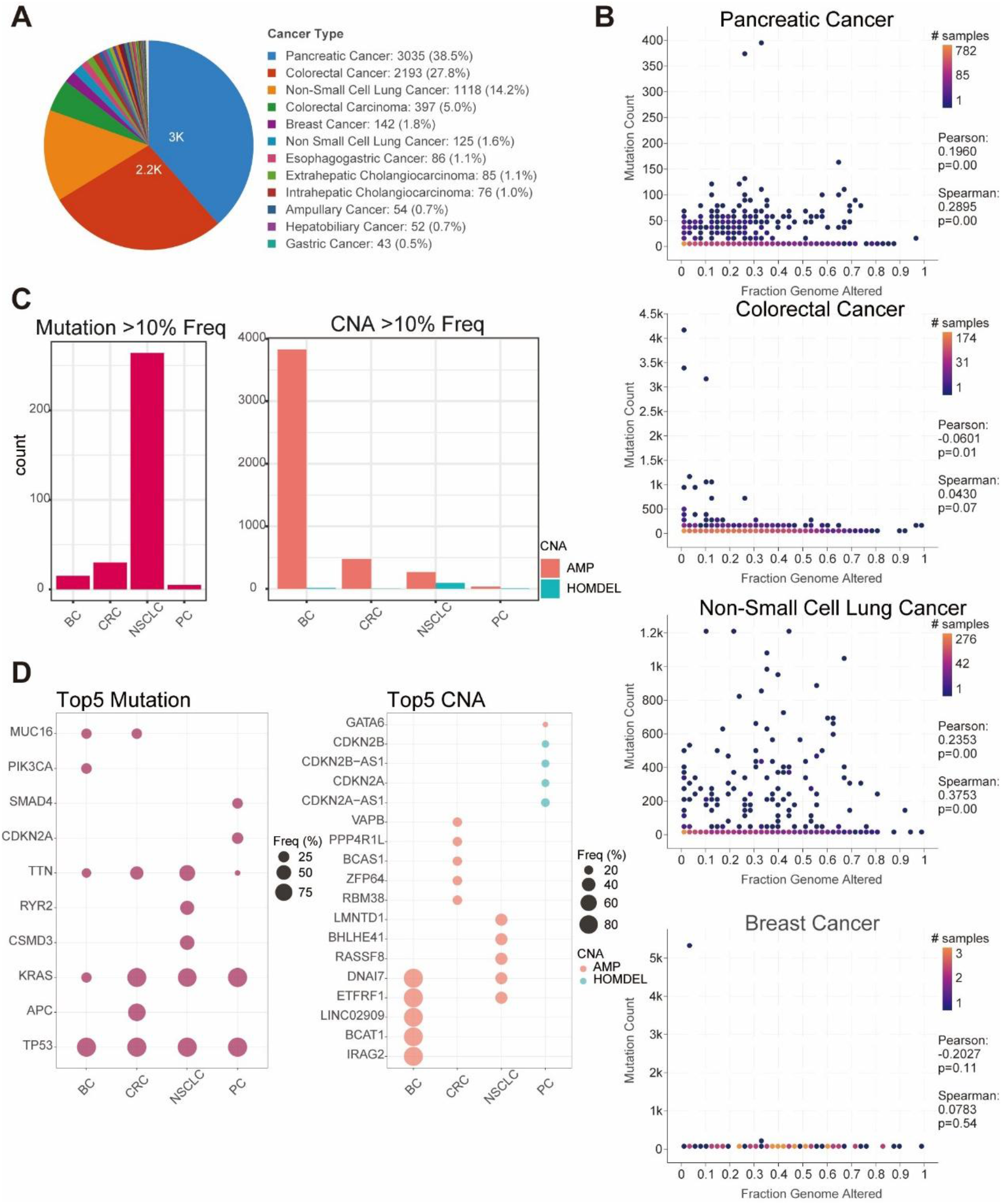
Genetic characteristics of double-positive samples across cancer types. **(A)** Pie chart showing the distribution of double-positive (*KRAS*^altered^/*TP53*^altered^) samples across cancer types. **(B)** Tumor mutational burden (TMB) in four cancer types. **(C)** Bar plot showing the number of high-frequency (>10%) mutations and CNAs across cancer types. **(D)** Bubble plot illustrating the top five co-occurring mutated and CNA genes in four representative cancer types.

**Supplementary Figure 3.**
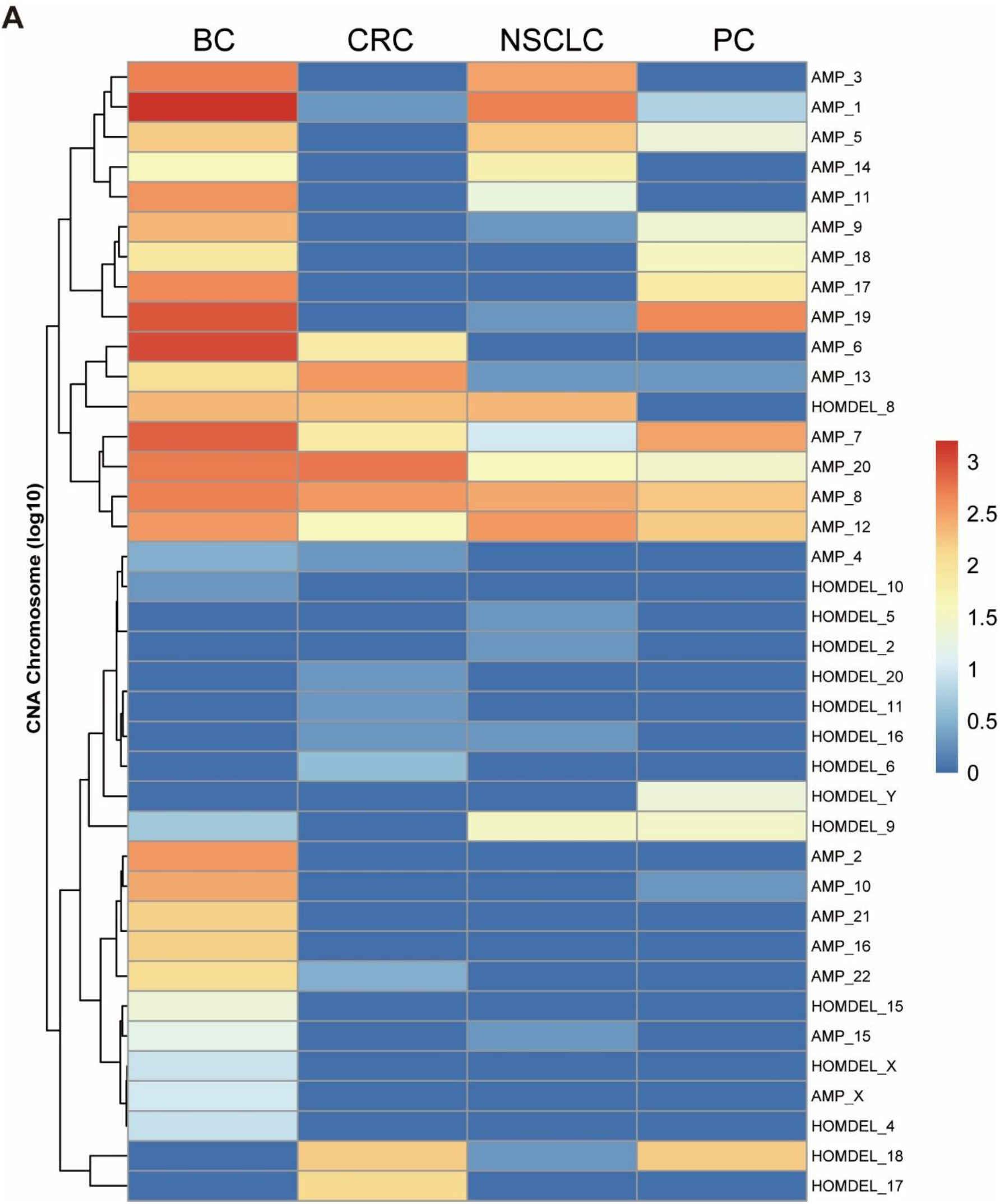
Chromosomal distribution of copy number alterations. Heatmap showing the log-transformed count of CNA across chromosomes in four representative cancer types.

Specialized statistical analysis was conducted on high-frequency CNA and gene mutation events with a proportion exceeding 10%. The results uncovered starkly distinct distribution preferences for mutations and CNAs across cancer types: NSCLC samples were mainly characterized by the enrichment of various high-frequency point mutations, while breast cancer samples were dominated by large-scale, high-frequency CNA events (Figure 4C). Notably, PC samples were highly accompanied by *CDKN2A* gene mutations and CNAs, whereas CRC samples featured high-frequency *APC* mutations as the hallmark concurrent genetic event, revealing completely distinct genetic signatures between the two cancer types (Figure 4D). Following precise mapping of CNA events to their corresponding chromosomal locations, it was evident that breast cancer exhibited the broadest range of chromosomal copy number alterations and involved the highest number of chromosomes among all enrolled cancer types. The remaining included cancer types each displayed unique and individualized chromosomal CNA profiles with no unifying pattern (Supplemental Figure 3).

Collectively, the comprehensive analysis highlighted that *KRAS*^*altered*^/*TP53*^*altered*^ patients exhibited striking cancer type-specific enrichment, predominantly clustering in PC, CRC and NSCLC. Among these, NSCLC presented a highly heterogeneous mutational spectrum with no single dominant mutational subtype; PC were characterized by frequent concomitant *CDKN2A* alterations, including both gene mutations and CNAs; while CRC cases were hallmarked by high-frequency *APC* mutations as the core concurrent genetic event. Overall, the background genetic landscapes of *KRAS*^*altered*^/*TP53*^*altered*^ patients varied drastically across distinct cancer types, with highly unique and cancer-specific genomic signatures.

Based on the above cancer-specific concomitant genetic alteration profiles, this study further performed stratified prognostic analysis to explore the associations between distinct background genetic features and clinical outcomes. In PC, *KRAS*^*altered*^/*TP53*^*altered*^ patients with concurrent *CDKN2A* alterations exhibited significantly shorter OS than *KRAS*^*altered*^/*TP53*^*altered*^ patients without *CDKN2A* co-alterations. Conversely, in CRC, *KRAS*^*altered*^/*TP53*^*altered*^ patients harboring concomitant *APC* mutations had significantly prolonged OS compared with their counterparts without *APC* mutations (Figure 5).

**Figure 5.**
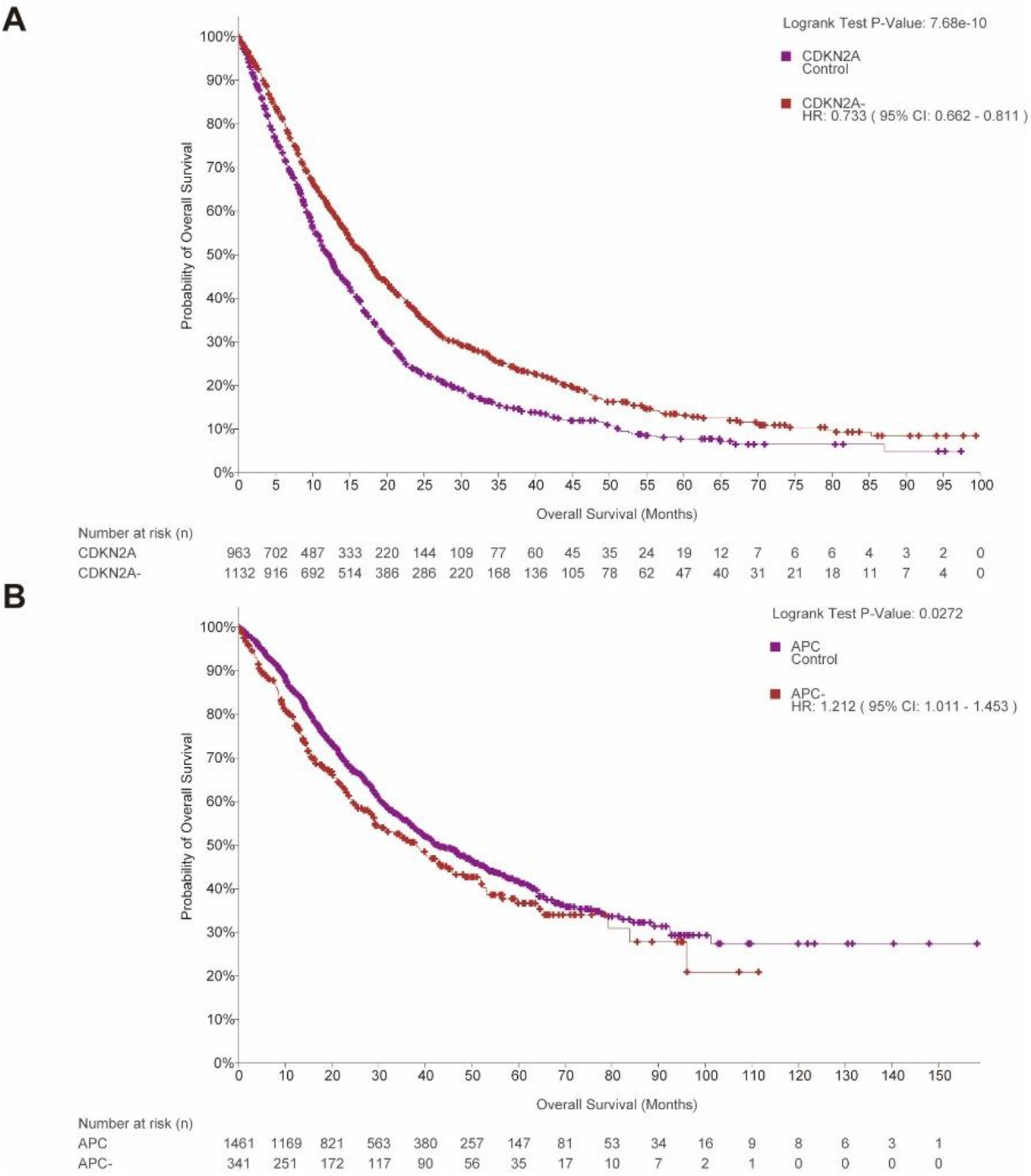
Prognostic impact of co-occurring genetic events in double-positive samples. Overall survival analysis of *KRAS*^altered^/*TP53*^altered^ pancreatic adenocarcinoma patients stratified by *CDKN2A* alteration status. **(B)** Overall survival analysis of *KRAS*^altered^/*TP53*^altered^ colorectal cancer patients stratified by *APC* mutation status.

## Discussion

By integrating pan-cancer cohorts, this study systematically elucidated the complexity and cancer-type specificity of *TP53* and *KRAS* alterations in prognostic evaluation. Although these two genes frequently exert synergistic effects during tumorigenesis, their associated clinical outcomes exhibit completely opposite trends depending on the distinct cancer-type backgrounds. Notably, this finding provides a crucial revision to the conventional perception that dual-gene alterations are uniformly equivalent to poor prognosis.

Our results demonstrate that concurrent TP53/*KRAS* alterations are associated with the poorest overall survival in PC, CRC, and ampullary carcinoma (Figure 1A–C), consistent with previous reports. In PC, dual mutations synergistically activate epithelial-mesenchymal transition (EMT), promote tumor dedifferentiation, and accelerate metastasis^28,29^, while frequent co-occurring *CDKN2A* alterations (Figure 4D) further exacerbate the malignant phenotype^30^. In CRC, dual alterations often co-exist with *APC* mutations (Figure 4D); however, their prognostic significance remains controversial. In our study, concurrent *APC* mutations were associated with longer survival in *KRAS*^*altered*^/*TP53*^*altered*^ CRC patients (Figure 5), suggesting that *APC* mutations may partially counteract the oncogenic effects of KRAS/TP53 by restraining aberrant Wnt pathway activation^31^.

In stark contrast, GC patients with dual alterations exhibited the longest overall survival (Figure 1E). This phenomenon may be attributable to the predominance of CNA in *KRAS* alterations in GC (Figure 2A) and the high frequency of microsatellite instability (MSI)^32^. Notably, the enrichment of *KRAS* G13D mutations in GC (Figure 2C)—a subtype previously reported to exhibit relatively indolent behavior in CRC^33^—further supports the profound impact of cancer-specific contexts on gene function.

We observed striking cancer-type distribution preferences for *KRAS* mutation subtypes (Figure 2C), likely reflecting tissue-specific mutational spectra and selective pressures. G12D/G12V mutations dominate in PC and CRC, both of which exhibit intermediate biochemical activity^34^, sufficient to drive transformation while avoiding excessive oncogenic stress. In contrast, G12C mutations predominate in non-small cell lung cancer (NSCLC), featuring a unique cysteine residue that has enabled the development of targeted inhibitors^35^. GC displays a distinct pattern characterized by *KRAS* G13D mutations and CNA, which may contribute to its relatively mild clinical course in this context.

Notably, *KRAS* CNA accounts for a substantial proportion of alterations in GC and NSCLC (Figure 2A), yet its prognostic significance differs from that of mutations (Figure 3B). Previous studies have shown that *KRAS* amplification correlates with aggressive phenotypes in EMC^36^ while potentially driving distinct transcriptional programs through dosage effects in GC^37,38^. These findings highlight that conflating CNA with mutations may obscure genuine biological differences. The ACRG dataset yielded differing prognostic outcomes, which may be attributable to ethnic differences, suggesting that larger sample sizes are needed to draw definitive conclusions.

In summary, this study systematically demonstrates that the prognostic value of TP53 and *KRAS* alterations is cancer-type specific, driven by differences in mutation subtype distribution, CNA patterns, co-occurring genetic events, and tumor microenvironment characteristics. These findings suggest that in clinical practice, dual-gene alterations should not be simplistically equated with poor prognosis; instead, assessment must incorporate cancer-specific contexts. Future studies integrating multi-omics data to develop cancer-type-specific prognostic models are warranted to inform precision treatment decisions. Furthermore, while targeted therapies for specific *KRAS* mutation subtypes, such as G12C inhibitors in NSCLC, have achieved breakthroughs^35^, effective strategies for G12D/G12V mutations—which predominate in PC—remain an urgent unmet need.

## Materials and Methods

### 1. Data Sources and Study Cohorts

This study utilized two complementary datasets for comprehensive genomic and prognostic analyses.

#### Multi-cohort dataset

To investigate the prognostic significance of *TP53* and *KRAS* alterations across different cancer types, we first compiled a large-scale multi-cohort dataset from the cBioPortal for Cancer Genomics (https://www.cbioportal.org/). We included studies with available mutation, copy number alteration (CNA), and survival data. The following cancer types were included based on sample size adequacy and data completeness: pancreatic adenocarcinoma (PC), colorectal cancer (CRC), ampullary carcinoma, non-small cell lung cancer (NSCLC), gastric cancer (GC), appendiceal adenocarcinoma, endometrial carcinoma (EMC), and cholangiocarcinoma. For each cancer type, we extracted patient-level genomic alteration data, including single nucleotide variants (SNVs), insertions/deletions (indels), and copy number alterations, as well as corresponding clinical follow-up information.

#### ACRG cohort (GSE62717)

To supplement gastric cancer analyses, we additionally included the Asian Cancer Research Group (ACRG) cohort, which contains copy number data and clinical annotations for 271 gastric cancer patients. The dataset was obtained from the Gene Expression Omnibus (GEO) under accession number GSE62717. For each sample, relative copy number values per gene were extracted. Clinical information, including overall survival, survival status, tumor stage, and Lauren histological subtype, was retrieved for prognostic analyses.

### 2. Definition of Genetic Alterations

For both the multi-cohort and TCGA datasets, genetic alterations were uniformly defined. Sequence variations included non-synonymous single nucleotide variants (SNVs), insertions, and deletions affecting *TP53* or *KRAS*. Copy number alterations were defined as homozygous deletions or amplifications, as annotated in the source datasets. Patients were classified into four mutually exclusive subgroups based on the alteration status of *TP53* and *KRAS*:

- *KRAS*^altered^/*TP53*^altered^ (dual altered)
- *KRAS*^altered^/*TP53*^wt^ (KRAS only)
- *KRAS*^wt^/*TP53*^altered^ (TP53 only)
- *KRAS*^wt^/*TP53*^wt^ (double wild-type)

Alterations with a frequency below 5% across the entire cohort were defined as low-frequency mutations for the purpose of mutation spectrum stratification.

### 3. Prognostic Analysis

Overall survival (OS) was the primary endpoint for prognostic assessment. For NSCLC patients, central nervous system progression-free survival (CNS PFS) was additionally evaluated. Survival curves were estimated using the Kaplan-Meier method, and differences among the four subgroups were compared using the log-rank test. For cancer types with sufficient sample size, univariate and multivariate Cox proportional hazards regression models were applied to assess the independent prognostic value of *TP53*/*KRAS* alteration status, adjusting for clinical covariates including age, sex, tumor stage, and treatment history where available.

### 6. Statistical Analysis

All statistical analyses were performed using R software (version 4.1.0) and the associated packages: *survival* and *survminer* for survival analysis, *ggplot2* for visualization, and *ComplexHeatmap* for heatmap generation. For comparisons between groups, the chi-square test or Fisher’s exact test was used for categorical variables, and the Mann-Whitney U test or Kruskal-Wallis test was used for continuous variables. A two-sided *P* < 0.05 was considered statistically significant, with adjustments for multiple comparisons applied where indicated using the Benjamini-Hochberg method.

## Data Availability

All data produced are available online at

